# AWARENESS AND USE OF MEDIA CHANNELS FOR INFORMATION DISSEMINATION ON WASTE DISPOSAL BY ENVIRONMENTAL HEALTH WORKERS FOR HYGIENIC LIFESTYLE. A STUDY OF MINNA, NIGERIA

**DOI:** 10.1101/2025.07.18.25331709

**Authors:** Oluwatosin Daniel Akobe, Amina Badaru Yusuf

## Abstract

The purpose of this study was to determine the Awareness and Use of Media Channels for Information Dissemination on Waste Disposal by Environmental Health Workers for Hygienic Lifestyle. A Study of Minna, Nigeria. Survey and content analysis research methods was be adopted for this study. The results obtained from the questionnaire, interviews and programmes schedule was analysed as part of discussion of findings. The total population of the study comprises 150 environmental health workers selected from four (4) departments that are made up of: waste management department, environmental monitoring, conservation department and environmental health. Based on the result, it is safe to say that the use of environmental health information has improved the hygienic lifestyle of the populace in Minna and tis has in turn improved the overall quality of the environment. It is observed that the access and dissemination of environmental health information has greatly decreased the outbreak of diseases among the populace which has positively impacted the reduction of cholera among the populace in Minna. However, there is still more work to be done by the environmental health workers. It was observed in the study there were inadequate and insufficient information dissemination by the environmental health workers on environmental issues in general and solid waste in particular. Attitudes towards solid waste management were poor and insufficient for sustainable environmental development.

## Introduction

The Management of solid waste is an enormous challenge in developing countries all over the world due to factors like: poverty, population explosion and urbanisation. The management is also ineffective and underfunded by governments (Adewale, 2011). A sustainable system in place for handling waste is an acute need within rural settlements. This is because many of these small towns are growing fast and increasing human populations will lead to production of more waste. The generation of Municipal Solid Waste (MSW) has been rapid, while the capacity to collect and safely dispose of the material has been on a general decline. Today, municipal solid wastes are getting disposed in open and illegal dump sites which is due to improper environmental pollution control and monitoring (Akobe et al., 2025).

Open dumpsite approach as solid waste disposal method is a primitive stage of solid waste management in many parts of the world. It is one of the most poorly rendered services by municipal authorities in developing countries as the systems applied are unscientific, outdated and inefficient. Solid waste disposal sites are found both within and on the outskirts of developing urban cities. With increase in the global population and the rising demand for food and other essentials, there has been a rise in the amount of waste being generated daily by each household (Foday et al., 2013). This waste is ultimately thrown into municipal disposal sites and due to poor and ineffective management, the dumpsites turn to sources of environmental and health hazards to people living in the vicinity of such dumps. One of the main aspects of concern is the pollution caused to the earth; be it land, air and water. According to Nguyen et al. (2011) many cities in developing countries face serious environmental degradation and health risks due to the weakly developed municipal solid waste management system. Several studies have been conducted in order to examine the health and environmental effects arising from waste dumps. Such studies showed that a link exists between the two (Aatamila et al., 2010). The ever-increasing consumption of resources results in huge amounts of solid wastes from industrial and domestic activities, which pose significant threats to human health (Foday et al., 2013).

However, the ills of inappropriately disposed municipal solid wastes are quite numerous to be mentioned. Health deterioration, accidents, flood occurrences, and environmental pressures are just a few of the negative effects. In many developing countries, solid waste disposal sites are found on the outskirts of urban areas. These areas become children’s sources of contamination due to the incubation and proliferation of flies, mosquitoes, and rodents. They, in turn, are disease transmitters that affect population’s health, which has its organic defences in a formative and creative state. The said situation produces gastrointestinal, dermatological, respiratory, genetic, and several other kinds of infectious diseases (Foday et al., 2013; Salam, 2010). Open dumpsites in developing urban cities involve indiscriminate disposal of waste. They are uncontrolled and therefore pose major health threats which affect the landscape of urban cities. Wastes that are not managed properly, especially solid waste from households and the community, are a serious health hazard and lead to the spread of infectious diseases. It is also evident that unattended wastes lying around attract flies, rats, and other creatures that, in turn, spread diseases. Normally, it is the wet waste that decomposes and releases a bad odour. The bad odour affects the people settled next to the dumpsite, which shows that the dumpsites have serious effects to people settled around or next to them (Omang, et al. 2021).

### Statement of Problem

Solid waste management and indiscriminate disposal of waste is one of the major problems faced by city planners in developing countries due to poor planning, increased urbanization and inadequate resources (Ogunbode, et al. 2021). Recently, there have been growing concerns about the environmental and health effects associated with the generation of solid waste as well as the increasing cost of municipal solid waste management. Improper waste disposal can result in environmental health hazards and negative impact on the environment in general. In major cities of Nigeria most especially Minna, the disposal of solid wastes within the last few decades have posed major environmental and public health issues as the majority of open dumpsites which are initially located in the outskirts are now within the heart of the city as a result of urbanisation and migration. Solid waste management is on a downward spiral in Nigeria, with most communities especially within city centres not benefitting from the municipal waste disposal services (Agbunu, et al., 2020).

In some parts of Nigeria, it is common to see refuse being dumped along major roads and highways constituting nuisance in those locations. This has become an eyesore as major streets have been turned into refuse dumps with ugly mountains of waste causing serious traffic problems (Olalekan et al., 2021). Asides the aesthetic issues caused by these improper disposal methods, the health and environmental implications cannot be ignored. The dumpsite is an ideal breeding ground for disease vectors such as rats and mosquitoes which present serious health issues to nearby household residents. Such unsanitary environment is a predisposing factor for the spread of diseases/infections like malaria, dengue fever, typhoid, tetanus, cholera, eczema and dysentery. Recently the emphasis has been directed towards participatory approaches in solid waste management in most developing countries. This involves participation of the concerned actors at various levels to enhance co-operation (Ogunyemi et al., 2023).

### Research Objectives

The specific objectives are to:

1. Determine the level of awareness by environmental health workers on the use of media channels for disseminating environmental information on waste disposal for hygienic lifestyle in Minna, Nigeria.
2. Identify the various media channels used for disseminating environmental information for effective waste disposal in Minna, Nigeria.
3. Ascertain the impact of information dissemination on waste disposal on the health of the populace for hygienic lifestyle in Minna, Nigeria.

### Literature Review

Studies have shown that information has helped to promote environmental behaviour. Relevant information can help individuals to understand the interaction between resources (natural) and the environment. It is observed that greater knowledge of environmental principles, attitudes and theories of waste reduction through access and use of information can enhance individual’s ability to participate in solid waste management. Information is an integral part of environmental management because it is central to every human activity and as such, would be used in coordinating the resources for a ‘synergistic’ approach to management of the environment (Ridayani et al., 2022).

Therefore, environment information consists of all forms of information to keep the public enlightened about and aware of environmental issues and trends. It is based on this reason that the role of information in solid waste management becomes imperative. These roles are to raise awareness in environmental issues and it can be used to promote responsible environmental behaviour, especially for solid waste management (Konstantinidou et al., 2024). In addition, it enables government and its agencies to know areas where solid waste management needs serious and urgent attention. In the same vein, Macawile and Sia Su (2009) believe that a conscious effort through information dissemination is needed to “incorporate the interests of both the community leaders and the public in understanding their roles, relationships and contributions through their perceptions and attitudes as all are recognised as important stakeholders in attaining a sustainable environmentally oriented effort”. In essence, communication is one of the vital ways by which people in any given environment could relate. The essence of environment which deals with living together of all organisms in the environment is only possible and realisable through the use of communication (Zhang et al., 2022).

Information dissemination in this study goes beyond mere sharing of information, but as a way of influencing beliefs, views, perceptions and to induce behavioural or attitudinal change. The public holds the media in high esteem in terms of information and enlightenment. McQuail (2010) believes that whenever the media exert influence, they also cause change. Information therefore, can be a potent force to influence public perceptions on various issues of life. Alabi (2010) noted that attitudes and predispositions are at work before and during exposure to information, and they in fact largely determine the use of information, how we interpret the contents and the effect which information has upon us. In other words, “messages received from the media affect our thoughts and beliefs formation as well as responses to attitude” (Alabi, 2010).

Information through effective communication can therefore be used to influence people’s dispositions for a friendly environment. Although, mass communication messages may not change existing deep-rooted attitudes but may rather influence it. The ultimate goal is to activate public efforts towards behavioural change in environmental management. Behavioural change for environmental management may sometimes require consistent and systematic applications or activities to achieve desired goals. This may be achieved through public enlightenment campaign by environmental health workers. A campaign is the planning and coordination of series of consistent activities aimed at achieving a central objective. In the same vein, McQuail (2010) defines a campaign as the planned attempt to influence public opinion, behaviour, attitudes and knowledge on behalf of some cause, person, institution or topic, using different media over a specific period of time. Public campaigns are usually directed towards socially approved goals. In this wise, environmental communication campaign entails the adoption of specific steps towards an environmental objective. The essence is to bring behavioural change, and inculcate an environmentally friendly attitude or culture in people.

According to Ogbu, (2025), environmental campaigns could be pursued using different media like radio and television. Radio, for example, is believed to be the most effective, popular and credible medium for reaching a large and heterogeneous audience. It is relatively cheap, available and accessible. It can be powered by battery, requiring insignificant literacy level to comprehend. Radio remains the most potent and effective environmental communication tool for reaching a vast range of audience in developing nations like Nigeria (Yusuf & Isiaka, 2025).

In the same development, television is believed to make the most impact on the audience. This is because of its audio-visual advantage. It leaves a lasting impression in the minds of the audience (Adebayo et al., 2021). Television can reach diverse people simultaneously and provide opportunity for a message to be demonstrated in images or pictures. In a study of inhabitants’ perception on domestic waste disposal in Ijebu Ode, Southwest Nigeria, Banjo, Adebambo and Dairo (2009) result showed that radio and television were the most available (93% and 96% respectively), the most easily accessed (70% and 73% respectively) and the most effective sources of environmental information (61% and 64% respectively). Their study showed the effectiveness of the mass media, particularly the radio and television in creating awareness about public health and environmental issues. Radio and television are often associated with their wide geographical coverage and the relatively cheap cost of acquiring and using them in contrast to the print media (Banjo et al. 2009). Both media (radio and television) are effective environmental communication tools which could be used to raise public awareness and consciousness towards environmental concerns with a high degree of effectiveness.

Broadcast media enlightenment campaigns on solid waste management can come in any form of: radio jingles, television commercials (green advertising), main news bulletin, public service announcement, health programmes and so on. It is instructive to know that the degree to which the broadcast media devote air time to environmental news also affect people’s attitudes towards the environment. As it were, heavy dependency and exposure to the media tend to shape people’s beliefs and perceptions about various issues of life. Aptly put, the degree of dependency on the media is a key variable that help to explain why audience’s beliefs, feelings or behaviours are altered (Peter, 2024).

In this wise, Idamah, (2015) asserted that environmental news is a potent force for responsible environmental behaviour. Individual exposure to a greater amount of environmental news is more likely to show concern with environmental management. Taken from above, much of contemporary environmental studies are predicated on the belief that human and non-human welfare are threatened by a growing array of human-induced environmental problems namely pollution, over-population, consumption of non-renewable biodiversity loss, ozone depletion, greenhouse warming and others. It is universally agreed that human behaviour has been and will continue to be, of central importance in identifying, understanding and dealing with such problems. Therefore, it can be taken that environmental behaviour is affected by the level of public awareness created by the mass media on environmental issues (Jege & Omopariola, 2025).

Most researchers only focus on the best practices to adopt for proper waste disposal without adequately looking at the perspective of proper information dissemination on the way to better practice effective waste disposal. This is the gap this research intends to fill. This research therefore will address the access, use and dissemination of information on waste disposal by environmental health workers for hygienic lifestyle of the populace.

### Methodology

Survey and content analysis research methods was be adopted for this study. Survey research describes variables like attitudes, opinion, values, beliefs, which leads to gathering of information about a group of people. Also, survey method allows the gathering of data from a large target population through the instrumentality of questionnaire and personal interviews. The survey method enabled the researcher to measure respondents’ opinions, feelings and attitudes to questions asked through a questionnaire. Content analysis is a study of printed materials in a systematic and quantitative way for the purpose of measuring variables. The results obtained from the questionnaire, interviews and programmes schedule was analysed as part of discussion of findings. The total population of the study comprises 150 environmental health workers selected from four (4) departments that are made up of: waste management department, environmental monitoring, conservation department and environmental health. The researcher adopted the entire population of the environmental health workers in Minna, Niger State. This is called total or complete enumeration or census. This is because the population size is manageable. Popoola (2011) maintained that a researcher can study or adopt the entire population when the population size is not too large.

## Findings

### Research Question 1

What is the level of awareness by environmental health workers on the use of media channels for disseminating environmental information on waste disposal for hygienic lifestyle in Minna, Nigeria?

Table 4.1 revealed that all the thirteen items (13) listed have a mean score greater than the benchmark mean of 2.50. This shows that environmental health workers are highly aware of the following information channels: radio, television, posters, newspapers, magazines, libraries and social media platforms as a means of information dissemination.

**Table4_1:**
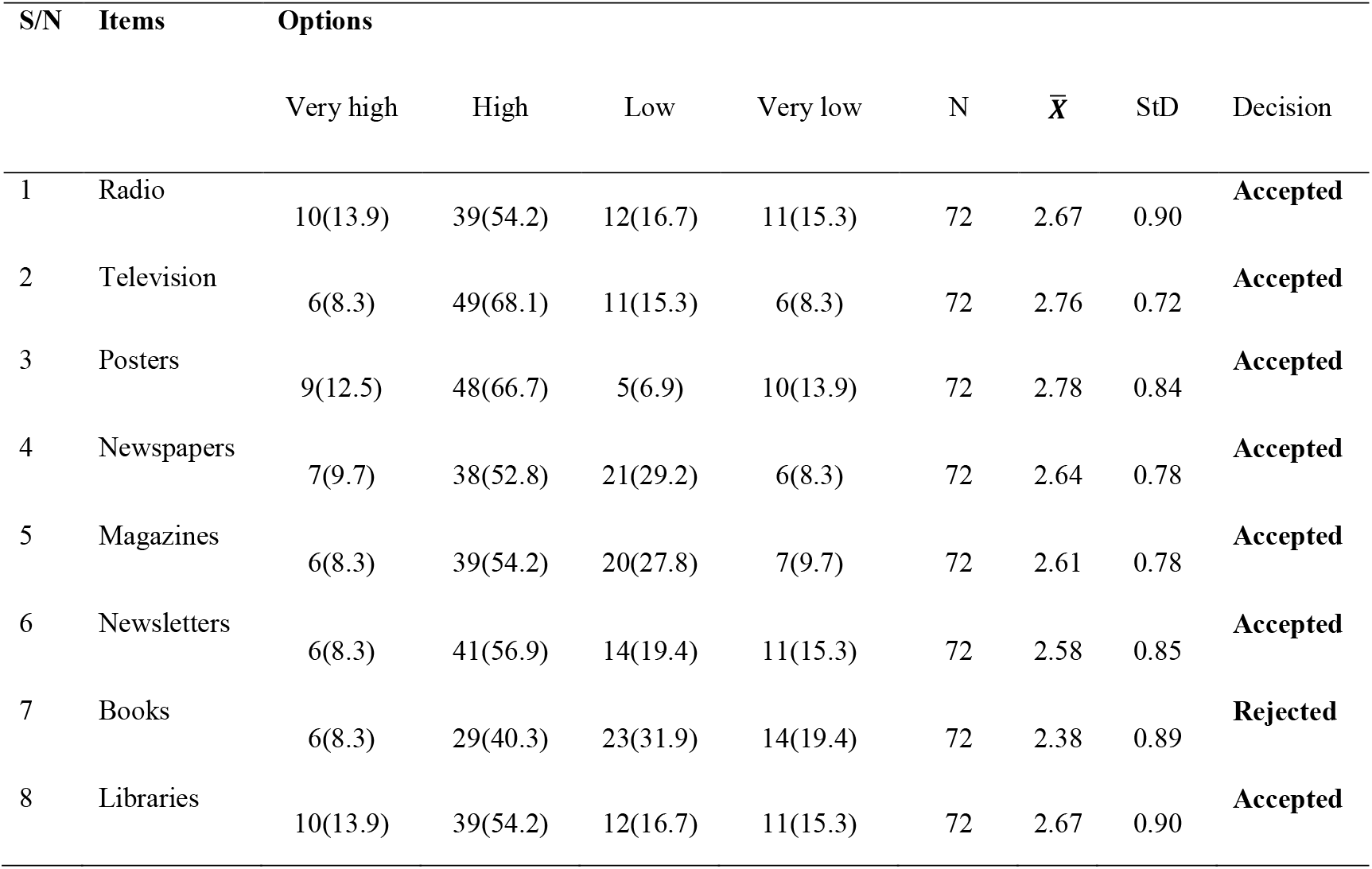
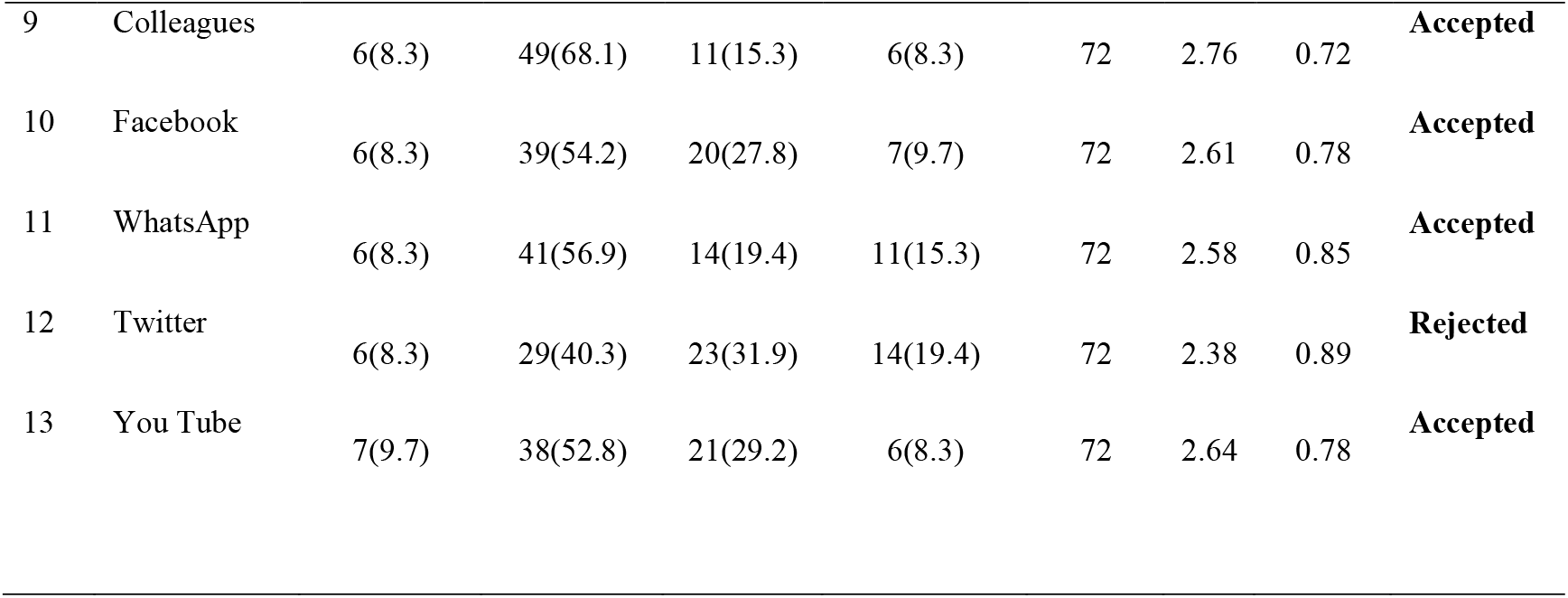
Level of Awareness of Information Channels.

### Research Question 2

What are the various media channels used for disseminating environmental information for effective waste disposal in Minna, Nigeria?

Table 4.2 revealed that all 13 items (13) listed have a mean score greater than the benchmark mean of 2.50. This means that the various media channels are used for disseminating environmental information for effective waste disposal in Minna, Nigeria.

**Table 4_2:**
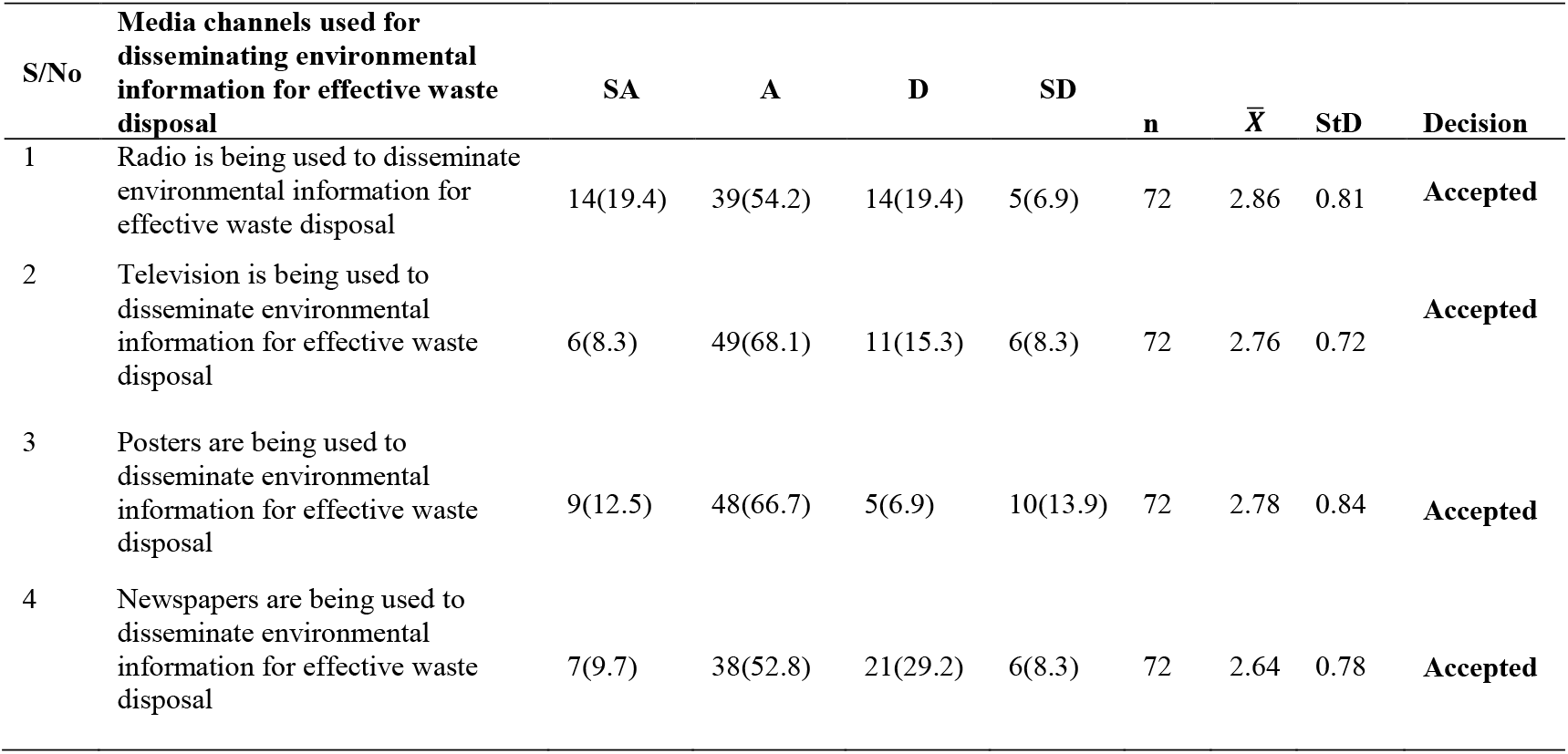
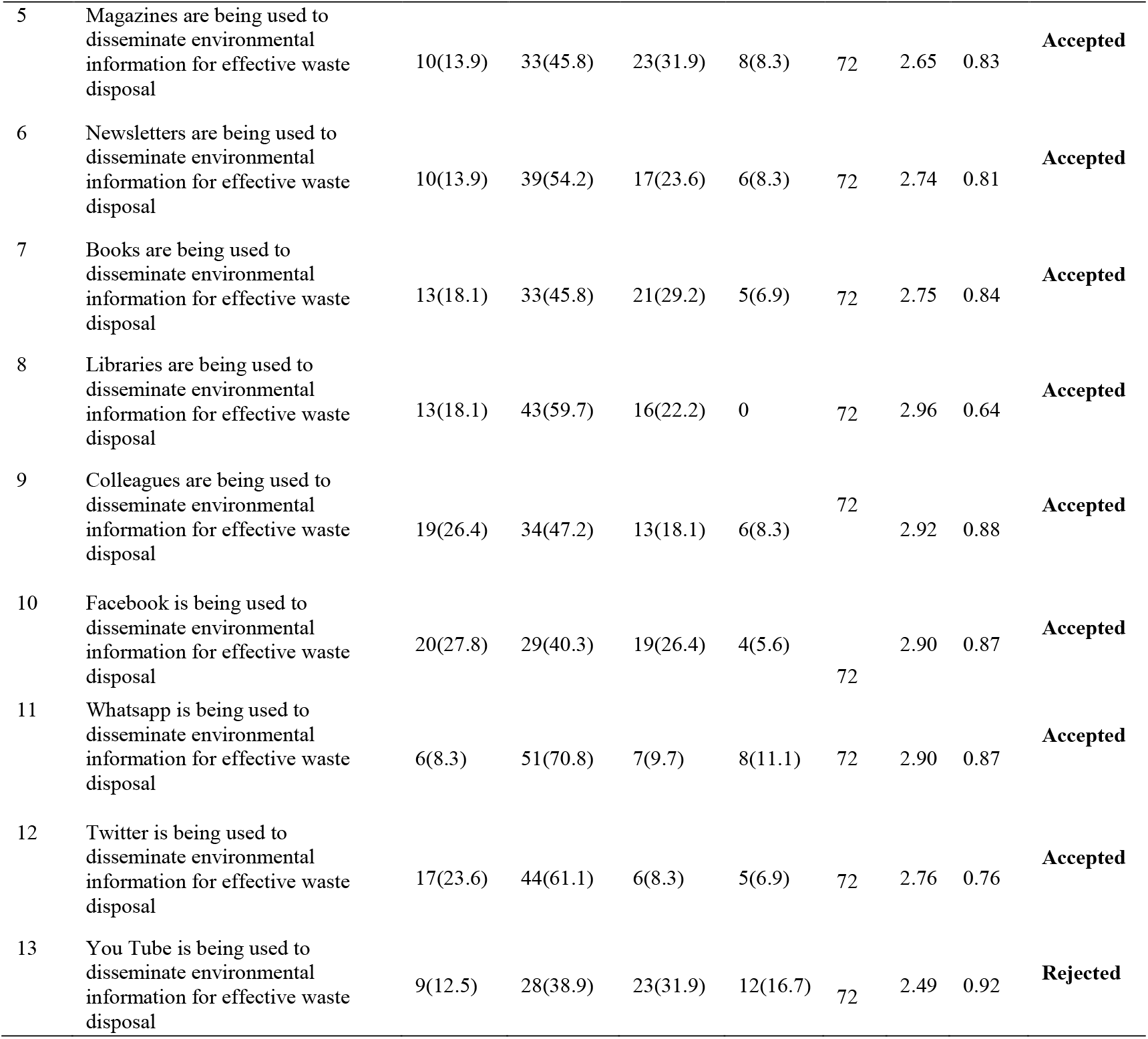
Impact of information dissemination on waste disposal on the health of the populace.

### Research Question 3

What is the impact of information dissemination on waste disposal on the health of the populace?

Table 4.3 revealed that eight (8) listed items have a mean score greater than the benchmark mean of 2.50. This means that Impact of information dissemination on waste disposal on the health of the populace has positively impacted the reduction of typhoid among the populace, it has influenced the once-a-month sanitary check which has greatly improved the health of the citizens, It has influenced the drastic reduction of the outbreak of diarrhoea among the populace and it has influenced the routine check of the environmental health workers which has greatly impacted on the quality of health of the populace.

**Table 4_3:**
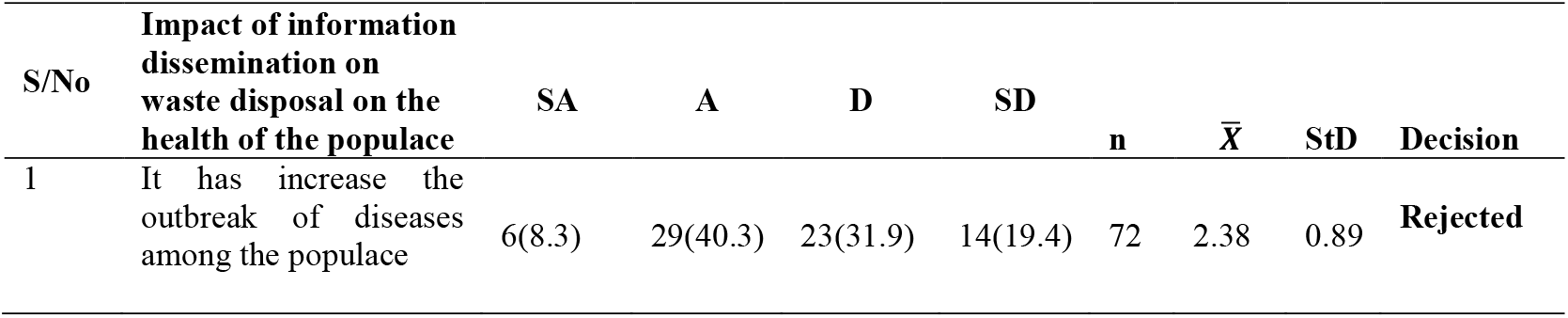
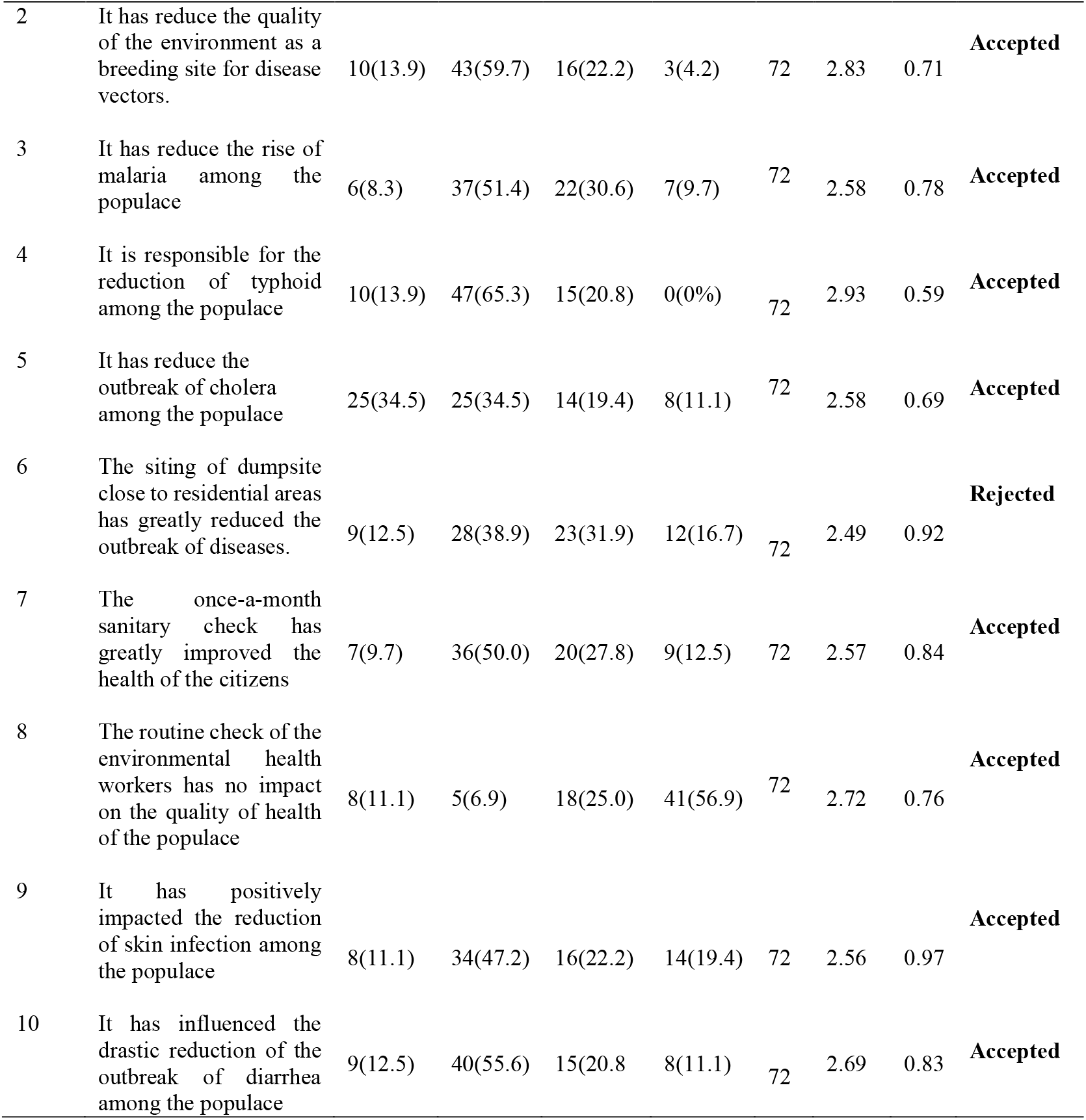
Impact of information dissemination on waste disposal on the health of the populace.

## Conclusion

Solid waste management is one of the greatest environmental challenges facing Nigeria. In order to inculcate positive attitude towards inhabitants, there is need to adopt enlightenment campaigns by environmental health workers. It is pertinent to conclude as follows: the access, use and dissemination of environmental information by health workers for hygienic lifestyle of the populace has great impact on the overall wellbeing of the people. However, there is still more work to be done by the environmental health workers. It was observed in the study there were inadequate and insufficient information dissemination by the environmental health workers on environmental issues in general and solid waste in particular. Attitudes towards solid waste management were poor and insufficient for sustainable environmental development. This is because many Nigerians still see the management of solid waste as largely the responsibilities of the local authorities. There is, therefore, the need for intensive enlightenment campaigns on solid waste management by environmental health workers for public behavioural change.

### Recommendations

The following recommendations are hereby made in accordance with the findings of the study:

1. There should be regular campaigns by environmental health workers in Minna on the environment which is essential for environmental management. This is because they help to shape social norms and values, influence people’s decision in ways that promote a more environmentally sustainable society.
2. Regular use of media channels to disseminate information on environment and health should be encouraged.

## Data Availability

All data produced in the present study are available upon reasonable request to the authors.

